# Effectiveness of Sinopharm’s BBIBP-CorV Booster Vaccination Against COVID-19-Related Severe and Critical Cases and death in Morocco During the Omicron Wave

**DOI:** 10.1101/2023.09.20.23295863

**Authors:** Jihane Belayachi, Abdelkader Mhayi, Hind Majidi, ElMostapha El Fahime, Redouane Abouqal

## Abstract

**Objective:** This study investigates the effectiveness of booster doses on the Omicron wave in Morocco against COVID-19 severe and critical hospitalizations and deaths;

**Participants/methods:** This study uses nationally representative data on COVID-19 from 15 December 2021 to 31 January 2022. To investigate the effectiveness of the inactivated COVID-19 vaccine BBIBP-CorV (Vero Cells) Sinopharm booster doses on the Omicron wave in morocco by using real-world data established from nationally representative statistics on COVID-19 cases, deaths and vaccinations.

**Statistical Analyses:** The screening method was used to estimate vaccine effectiveness against COVID-19 severe or critical hospitalization and COVID-19-related deaths. The data were grouped by, age subgroup, sex, week, and geographical area and were analyzed by using binary logistic regression with an offset for vaccine coverage.

**Results:** The overall sinopharm VE estimate is 89% (95% CI 85 to 92) effective in curbing COVID-19 death, and 81% (95% CI 78 to 84 in curbing COVID-19 severe critical hospitalization. Death-related VE estimate was 86% (95% CI 81 to 90) for patients aged 1Z65 years, 96% (95% CI 90 to 98) for those aged < 65 years, 95% (95% CI 88 to 98) in no risk factor patient was, 91% (95% CI 85 to 94) with 1 risk factor; 90% (95% CI 83 to 95) with 2 risk factors; 72% (95% CI 52 to 84) in patient with 3 risk factors and more. Severe critical hospitalization VE, estimate was 78% (95% CI 74 to 82) for patients aged 1Z65 years, 87% (95% CI 82 to 90) for those aged < 65 years, 86% (95% CI 80 to 90) in no risk factor patient was, 80% (95% CI 73 to 84) with 1 risk factor; 80% (95% CI 70 to 85) with 2 risk factors; 80% (95% CI 68 to 86) in patient with 3 risk factors and more.

**Conclusions:** Sinopharm Boosters are effective in increasing protection against Omicron variant-related COVID-19 death and severe critical hospitalization. The protection is reduced with older age and higher risk factors. These findings emphasize the importance of targeted vaccination strategies for different demographic groups and underscore the protective benefits of the third booster Sinopharm vaccine.

## INTRODUCTION

The coronavirus disease 2019 (COVID-19), caused by severe acute respiratory syndrome coronavirus 2 (SARS-CoV-2), has placed a significant burden on the healthcare system. As of early 2023, the SARS-CoV-2 virus continues to circulate globally. As of August 2023, the World Health Organization (WHO) has reported 769,369,823 confirmed cases of COVID-19, including 6,954,336 deaths, with a total of 13,492,225,267 vaccine doses administered (1). Challenges in public health persist due to the serial emergence of SARS-CoV-2 variants (2).

In the current phase of the COVID-19 pandemic, it is crucial to conduct a comprehensive assessment of the effectiveness of vaccines against severe SARS-CoV-2 infections. In this context, it is essential to analyze the overall effectiveness of both initial and booster vaccine doses, considering the specific vaccine types used. This evaluation contributes to measuring the protection offered against various manifestations of COVID-19, with particular emphasis on investigating the potential impacts of emerging concerning variants, including the Omicron variant (B.1.1.529). This analysis examines changes in vaccine effectiveness against these variants and their implications for public health strategies, with specific concern for vulnerable populations such as the elderly, immunocompromised individuals, and those with chronic health conditions. The primary variants, including the Omicron variant (B.1.1.529), exhibit distinct characteristics related to virulence, transmissibility, and vaccine evasion (3). Research on immunogenicity suggests that vaccine effectiveness may decline over time; however, it remains unclear how this reduction translates to clinical vaccine effectiveness.

In a recent meta-analysis conducted by Wu et al., encompassing 68 studies on the long-term vaccine effectiveness of COVID-19 vaccines (mRNA and Non-Replicating Viral Vector) against infections, hospitalizations, and mortality, the authors noted significant decreases over time in vaccine effectiveness for SARS-CoV-2 infections, hospitalizations, and mortality for both the primary vaccine series and booster doses. The Omicron variant exhibited lower levels of vaccine effectiveness at baseline, with further reductions over time (4). Estimates of baseline vaccine effectiveness against hospitalizations in response to the Omicron variant were 71%, seven days after vaccination. The patterns of vaccine effectiveness change were similar for those vaccinated against the Omicron variant compared with the general data on any variant. The authors suggested that vaccine effectiveness findings align with data on immunogenicity, which indicates that the robust immunological response initially elicited by vaccination appears to diminish over time.

Regarding inactivated vaccines, a recent study by Huang et al. demonstrated that among vaccinated individuals, administering a third homologous dose, compared to receiving two doses of inactivated vaccine at least 181 days prior, was associated with a 60.5% reduction in the incidence of severe/critical illness and an 81.7% reduction in the incidence of COVID-19-related death. Booster vaccination provided the greatest protection associated with the intensity of control and prevention against COVID-19 epidemics. (5)

Our previous study examined the primary regimen vaccine effectiveness of BBIBP-CorV (Vero Cells) Sinopharm against severe or critical hospitalizations due to COVID-19. This study found that the Sinopharm vaccine is highly protective against serious SARS-CoV-2 infection under real-world conditions. Protection remains high and stable during the first three months following the primary dose but decreases slightly beyond the fourth month, especially in patients aged 60 years and older (6). The rapid increase in COVID-19 cases resulting from the Omicron variant in vaccinated populations has raised concerns about the effectiveness of booster doses of current vaccines against severe or critical cases requiring hospitalization, as well as against mortality.

Morocco has experienced three epidemic waves, each corresponding to distinct periods marked by the emergence of noteworthy SARS-CoV-2 variants. The first wave, occurring between February and May 2021, was characterized by the prevalence of the Alpha variant (B.1.1.7). The second wave, taking place from July to September 2021, witnessed the predominance of the Delta variant (B.1.617.2), accounting for 80% of infections. The third and most recent wave has been characterized by the widespread circulation of the Omicron variant. Since August 2021, only BNT162b2 (Pfizer-BioNTech) and BBIBP-CorV (Vero Cells) Sinopharm vaccines have been deployed for the third and fourth booster doses (figure 2).

Inactivated vaccines are under-studied in real-life conditions, despite being deployed in nearly 100 countries. The variations in vaccine effectiveness against the Omicron variant (B.1.1.529), especially for vulnerable populations, require further investigation Here, we present vaccine effectiveness based on data obtained during the Omicron wave in Morocco. As of now, there is a scarcity of studies assessing the effectiveness of a BBIBP-CorV (Vero Cells) Sinopharm booster dose against the Omicron variant in the adult population. (7,8).

Hence, this study aims to assess the effictiveness of the inactivated COVID-19 vaccine BBIBP-CorV (Vero Cells) Sinopharm booster shots during the Omicron wave in Morocco. This evaluation relies on real-world data derived from nationally representative statistics on COVID-19 cases, fatalities, and vaccinations. Consequently, we seek to estimate the vaccine’s effectiveness against severe or critical hospitalization due to COVID-19 and COVID-19-related deaths utilizing the screening method.

## METHODS

### Study design

We estimate the real-world vaccine effectiveness (VE) after administration of booster doses of inactivated COVID-19 vaccine BBIBP-CorV (Vero Cells) Sinopharm against COVID-19-associated severe or critical hospitalized cases; and covid 19 related death between week 51/2021 and week 9/2022, using screening method. Population vaccine effectiveness is defined as the reduction in disease risk among vaccinated versus unvaccinated persons in the population.

The screening method can be used to estimate Vaccine effectiveness if information is available on the vaccine uptake of the population and on the proportion of vaccinated among the infected patients. Then, “screening method,” approach, uses estimates of 1) the proportion of persons with disease who are vaccinated (cases) and 2) the proportion of persons in the population who are vaccinated (reference population) (9).

### Data collection : cases

#### Severe-critical hospitalizations and deaths COVID 19 surveillance system

Individual-level data collected on confirmed covid19 severe/critical hospitalizations; and death during the period from15 December 2021 to 31 January 2022, were identified.

Clinical data concerning COVID-19 severe or critical hospitalization, and death are registered in a dataset made for public health surveillance systems. Vaccination status (vaccine product received, number of doses, and administration dates) for all sampled cases were extracted from national register of vaccination (RNV) based on national identity number, the clinical information on COVID-19 were linked to to the database of vaccinations, thereby providing both the date and the brand of the first, second and third dose of vaccination – or the lack of thereof – for each infected person. Patients were classified as 1) unvaccinated not having received any dose from any vaccine, 2) Primary vaccinated : 14 days after receiving either the second of two recommended doses of a two-dose vaccine or a single dose of the Janssen vaccine, regardless of whether the person received a third booster dose 3) Booster dose : 14 days after receiving the third dose (10).

- All patients that met the following criteria were included in the study:
- Age ≥ 18 years
- Within the opportunity to receive a third dose of covid 19 vaccine.
- Hospitalization for severe or critical case
- rt-Pcr laboratory-confirmed covid 19 occurring within 14 days prior hospitalisation

Severe and critical COVID-19 was defined in accordance with the WHO’s criteria. The WHO classifies severe COVID-19 as an individual infected with SARS-CoV-2 who exhibits an oxygen saturation of less than 90% while breathing room air and/or a respiratory rate exceeding 30 breaths per minute in adults, or displays signs of severe respiratory distress. Critical COVID-19 is defined as an individual infected with SARS-CoV-2 who develops acute respiratory distress syndrome, sepsis, septic shock, or other conditions that would typically necessitate the administration of life-sustaining therapies, including mechanical ventilation (invasive or non-invasive) or vasopressor therapy (11).

The opportunity to receive a third dose of covid 19 vaccine was assessed by the time between primary vaccination (two dose) and hospitalization. Severe or critical hospitalization case 5 months and more from the primary vaccination is considered within opportunity to receive the third dose. Only patient receipt of Sinopharm vaccine Booster dose were included regardless of vaccine brand received for the primary vaccination.

were excluded patients receiving a pfizer third dose, and those with any dose of any COVID-19 vaccine received <14 days before hospitalization were excluded.

For each case, dataon age, sex, date of COVID 19 hospitalisation, survival statut, lenght of stay specific clinical risk group status (the presence or absence of chronic respiratory disease, chronic heart disease, chronic kidney disease, diabetes or HTA) was extracted from the clinical surveillance systems.

### Data collection: reference population

Population-level vaccination coverage was determined using deidentified person-level COVID-19 vaccination data.

Vaccination data were obtained from a common, unbiased electronic health record system. This dataset was based on a comprehensive and inclusive population-based list of target populations drawn up firstly for the entire population over the age of 17 based on a national identity card (NIC) (representing the general population from which the reported cases have emerged). This list is the basis of the national vaccination register (NVR). Vaccine receipt is associated with the registration of vaccine information in the NVR.For a given week, data on population vaccination coverage two weeks prior to the week of case hospitalisation or death was obtained including the period 15 december 2021 - 31 January 2022(inclusive) corresponding to the omicron wave.

Vaccination status was classified as described for hospitalized cases.

The combined weekly and cumulative Data on vaccine uptake,including the weekly number of first, second and third doses administered from each vaccine brand, stratified according to age groups (18-19, 20-24, 25-29, 30-34, 35-39, 40-44,45-49, 50-54, 55-59, 60-64, 65-69, 70-74, 75+ years of age) were obtained from theRNV. To adjust for major confounders, the appropriate vaccine coverage was matched to cases at an individual level based on age group (18-19, 20-24, 25-29, 30-34, 35-39, 40-44,45-49, 50-54, 55-59, 60-64, 65-69, 70-74, 75+ years of age) and week of onset.

### Ethical Considerations

This real-world study was pre-registered (https://osf.io/at3yf) on Open Science Framework. The study was approved by the Rabat local ethic committee review board for biomedical research at Mohammed V University (N/21). A waiver of informed consent was granted for the study. Authorization N° A-RS-638/2021 from the National Commission for the Protection of Personal Data (CNDP) was obtained.

### Statistical Analysis

This screening Method predicts the vaccine effectiveness against an outcome of a disease Y (COVID-19 related death and severe critical COVID-19 hospitalisation), for the vaccination status d (full or boosted vaccination, in aall types of vaccine), using the following variables calculated collectively:

- The proportion of the population vaccinated, PPV;
- The proportion of outcomes within the vaccinated population, PCV.

Vaccine effectiveness against two outcomes of infections was determined using this method, namely against total severe critical COVID-19 hospitalisation and covid 19 related death.

Vaccine effectiveness was calculated using the following formula:

[1-(PCV(1-PPV)] / [(1-PCV) PPV]

Screening method (SM), compares the vaccination coverage between reported cases and a reference group. This simple method was designed to be used as a rapid preliminary analysis when incidence and attack rate data are not yet available (12).

The data were grouped by, age subgroup, sex, week, and geographical area and were analysed by using binary logistic regression with an offset (incorporating expected PPV by third dose and Sinopharm vaccine) for vaccine coverage. When estimating vaccine effectiveness for third dose receipt, patient who had received one or two doses were excluded.

To obtain the adjusted VE estimates with 95% CIs, logistic regression was conducted with the outcome variable as the vaccination status of the case (PCV), and with an offset for the log-odds of the matched coverage.The same adjustment was included for population vaccination coverage.

We performed sensitivity analysis according to age group (65≥, <65 years) and risk factors count group (0 no risk factor, 1: presence of 1 risk factor, 2: presence of 2 risk factors, ≥3: presence of 3 risk factors or more) to estimate the COVID-19 related death and severe critical COVID-19 hospitalisation Vaccine effectiveness.

*P* values were calculated to test the significance of the difference within each subgroup. A *P* value less than 0.05 was considered significant. For all point estimates of vaccine effectiveness, we calculated 95% CIs.

## RESULTS

### Description of cases

During the study period, 1741cases were hospitalized for severe or critical Covid19, and 699 cases were eligible for inclusion in the study and encompasse the including criteria. The flowchart of the study is shown in Figure 1. Among hospitalized patients, 35.3% received third sinopharm booster dose, Median age was 70 years with 66.4% aged more than 65years old. A risk factor was absent in 27.2% of hospitalized patients. Mortality rate was 35.6% Baseline characteristics of severe critical covid19 Hospitalisation are summerized in Table 1.

**Figure 1:**
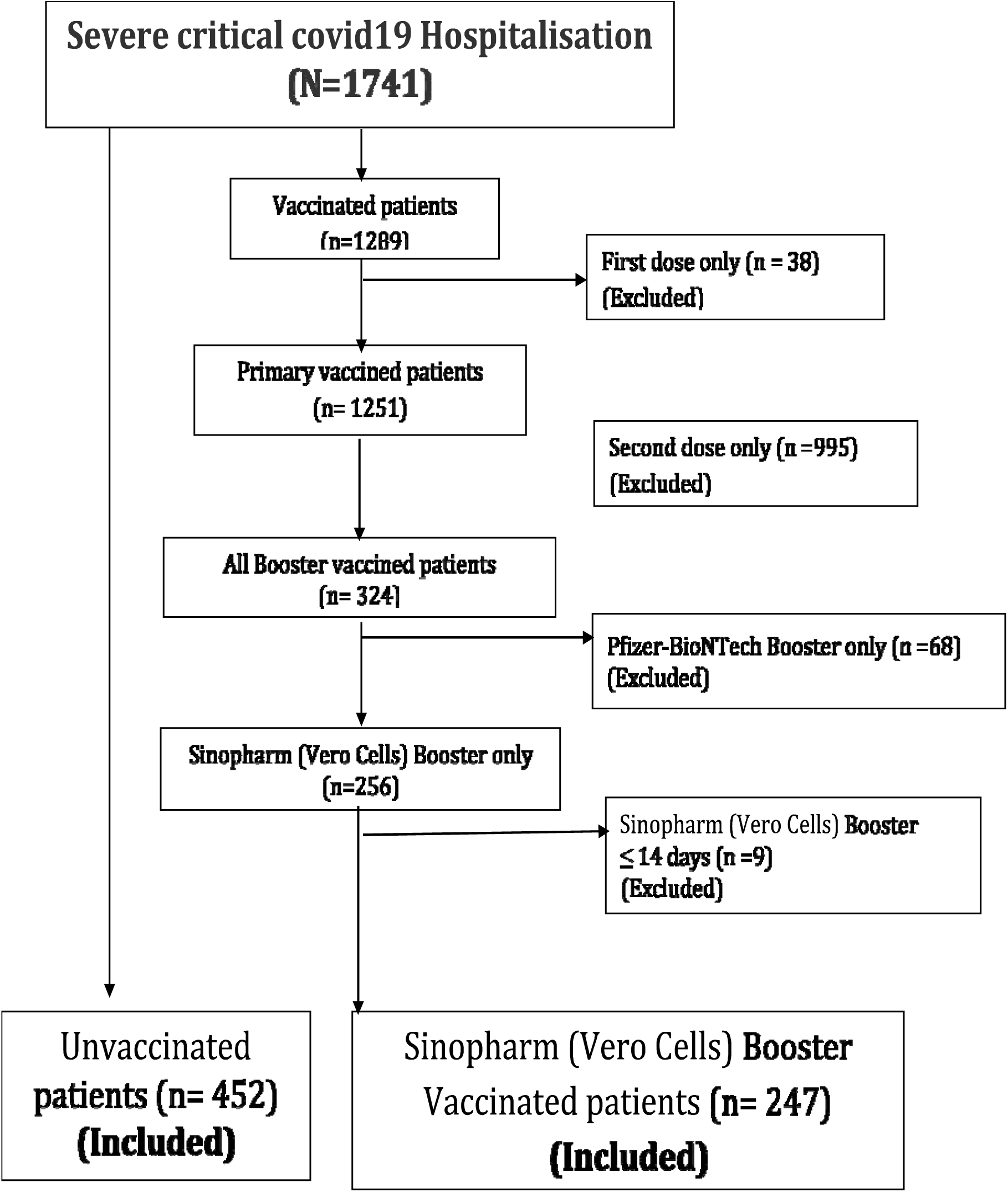
Flowchart of the study.

**Table 1:** Baseline characteristics of severe critical covid19 Hospitalisation.

|  | **Overall severe critical covid19 Hospitalisation during study period (N=1741)** | **Overall severe critical covid19 Hospitalisation included in the study (N=699)** |
| --- | --- | --- |
| **Sexe** |  |  |
| Female | 774 (44.5%) |  |
| Male | 967 (55.5%) |  |
| **Age median [IQR]** | 69 [ 60-79 ] | 70[61-80] |
| **Age subgroup** |  |  |
| ≤ 39 yrs | 149 (8.6%) | 64 (9.2%) |
| 40-59yrs | 278 (16.0%) | 89 (12.7%) |
| 60-64yrs | 201 (11.5%) | 82 (11.7%) |
| ≥ 65yrs | 1113 (63.9%) | 464 (66.4%) |
| **Risk factor count** |  |  |
| 0 | 481 (27.6%) | 190 (27.2%) |
| 1 | 673 (38.7%) | 263 (37.6%) |
| 2 | 347 (19.9%) | 142 (20.3%) |
| 3 | 240 (13.8%) | 104 (14.9%) |
| **comorb1** |  |  |
| 0 | 481 (27.6%) | 190 (27.2%) |
| 1 | 1260 (72.4%) | 509 (72.8%) |
| **Risk factor** |  |  |
| Diabetes | 553 (31.8%) | 227 (32.5%) |
| Hypertension | 598 (34.3%) | 271 (38.8%) |
| Chronic obstructive pulmonary disease and Other respiratory disease | 289 (16.6%) | 100 (14.3%) |
| cancer | 238 (13.7%) | 100 (14.3%) |
| Heartdisease | 327 (18.8%) | 147 (21.0%) |
| Chronickidneydisease | 167 (9.6%) | 60 (8.6%) |
| **statut_vacc** |  |  |
| unvaccinated | 452 (26.0%) | 452 (64.7%) |
| Only first dose | 38 (2.2%) |  |
| Primaryvaccinated | 927 (53.2%) |  |
| Third booster dose | 324 (18.6%) | 247 (35.3%) |
| **Survivalstatus** |  |  |
| 0 | 1178 (67.7%) | 452 (64.4%) |
| 1 | 563 (32.3%) | 247 (35.6%) |
| **Lenght of stay ( by days ) Mean (±SD)** | 6.5 ±4.6 | 6.4 ±4.7 |
| **Primaryvaccinationproduct** |  |  |
| VeroCells) Sinopharm | 457 (36.5%) | ----- |
| Pfizer-BioNTech | 5 (0.4%) | ----- |
| Oxford-AstraZeneca | 789 (63.1%) | ----- |
| **Third dose vaccine product** |  |  |
| VeroCellsSinopharm | 256 (79.0%) |  |
| Pfizer-BioNTech | 68 (21.0%) | ----- |

### VACCINE EFFECTIVENESS

#### COVID 19 Death related VE

The overall VE estimate was 89% (95% CI 85 to 92). VE estimate was 86% (95% CI 81 to 90) for patients aged 1Z65 years, and 96% (95% CI 90 to 98) for those aged < 65 years (figure3). According to risk factors count group, VE estimate in no risk factor patient was 95% (95% CI 88 to 98), 91% (95% CI 85 to 94) with 1 risk factor; 90% (95% CI 83 to 95) with 2 risk factors; 72% (95% CI 52 to 84) in patient with more than 3 risk factors (figure4).

#### COVID 19 Severe critical hospitalization VE

The overall VE estimate was 81% (95% CI 78 to 84). According to age group, VE estimate was 78% (95% CI 74 to 82) for patients aged 1Z65 years, compared to 87% (95% CI 82 to 90) for those aged < 65 years (figure3). According to risk factors count group, VE estimate in no risk factor patient was 86% (95% CI 80 to 90), 80% (95% CI 73 to 84) with 1 risk factor; 80% (95% CI 70 to 85) with 2 risk factors; 80% (95% CI 68 to 86) in patient with more than 3 risk factors (figure 4).

**Figure 2:**
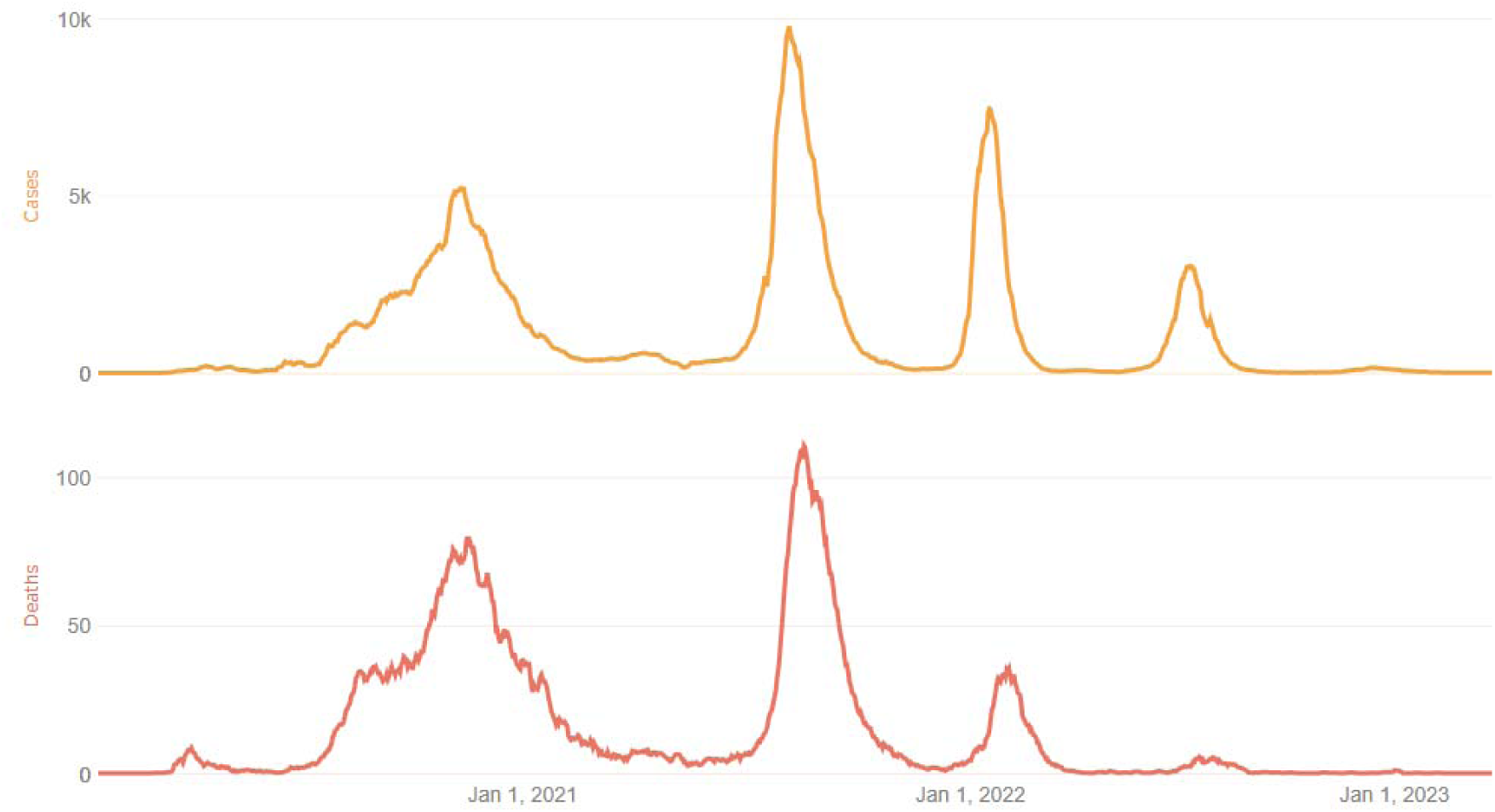
COVID 19 epidemic waves in MOROCCO

**Figure 3:**
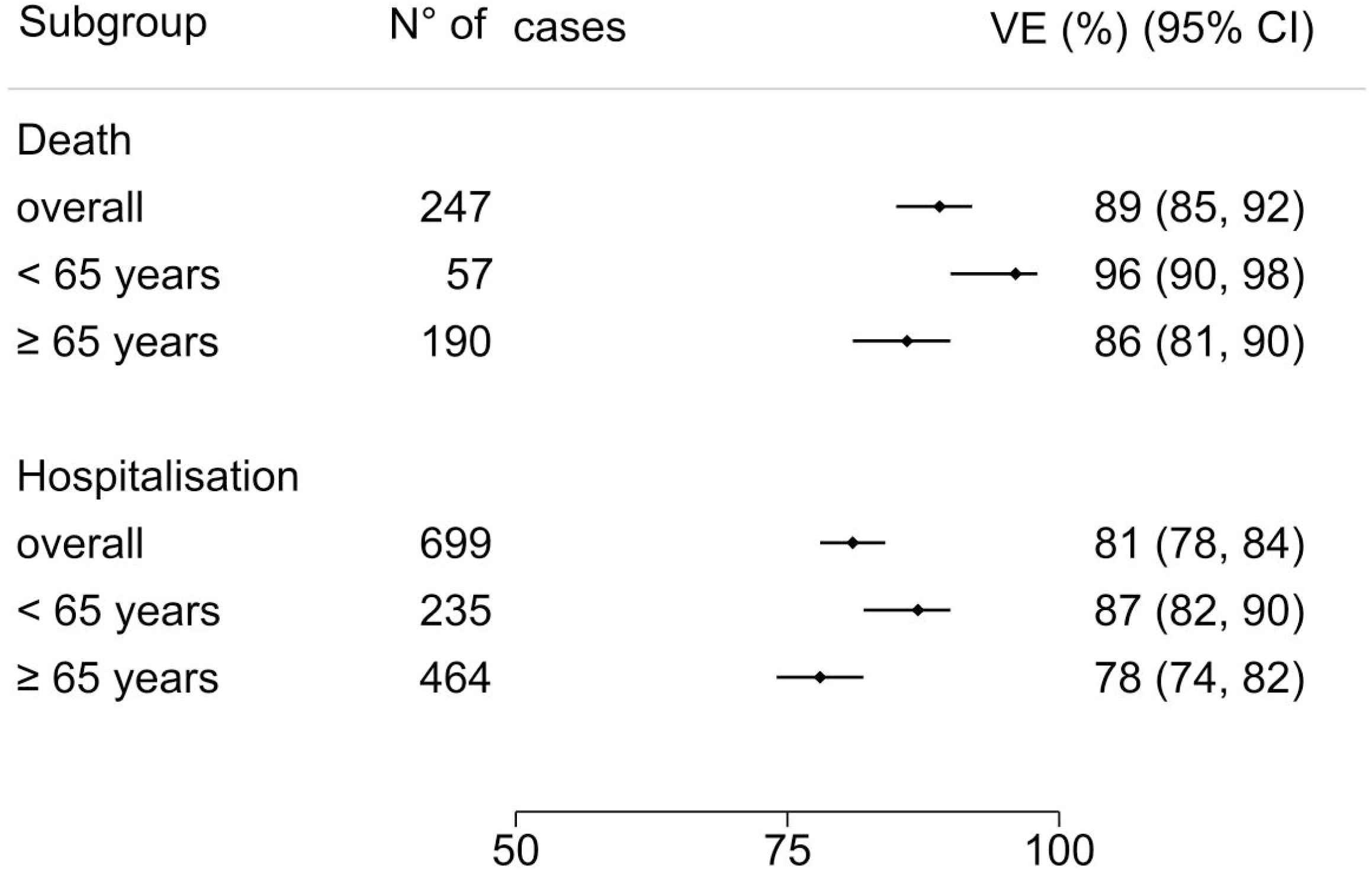
Covid 19 Related Death and hospitalization Vaccine effectiveness for age subgroups: lll65 years and < 65 years. Error bars indicate the %[CI95%]. The overall VE Covid 19 related death and related severe critical hospitalization was 89% (95% CI 85 to 92), and 81% (95% CI 78 to 84) respectively. In age subgroups, death VE estimate was 86% (95% CI 81 to 90) for patients aged lll65 years, and 96% (95% CI 90 to 98) for those aged < 65 years, and The severe critical hospitalization VE estimate was 78% (95% CI 74 to 82) for those aged lll65 years, and 87% (95% CI 82 to 90) for those aged < 65 years.

**Figure 4:**
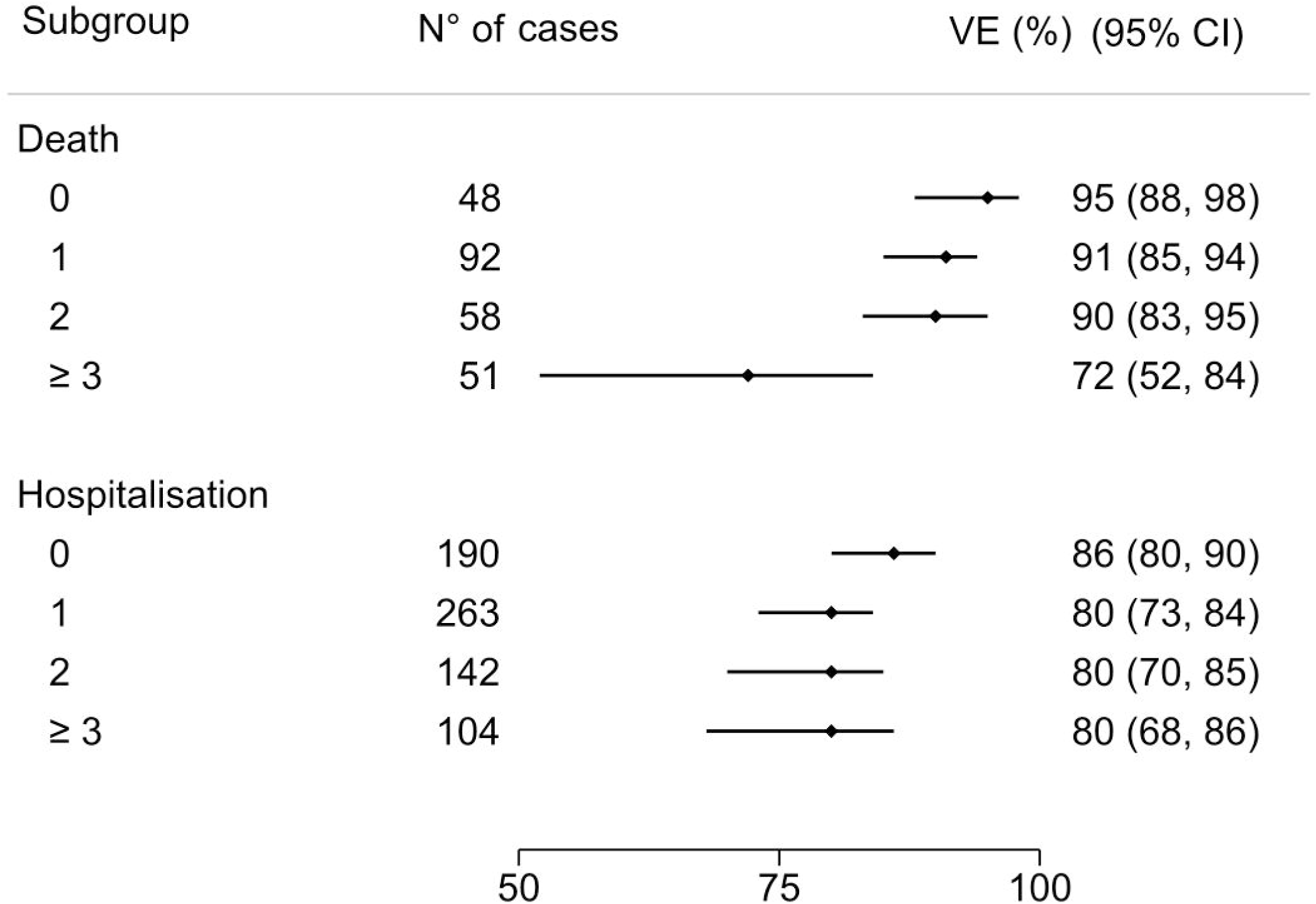
Covid 19 Related Death and hospitalization VE for risk factor count subgroups: Death VE estimate in no risk factor patient was 95% (95% CI 88 to 98), 91% (95% CI 85 to 94) with 1 risk factor; 90% (95% CI 83 to 95) with 2 risk factors; 72% (95% CI 52 to 84) in patient with more than 3 risk factors The VE estimate among severe critical hospitalization in no risk factor patient was 86% (95% CI 80 to 90), 80% (95% CI 73 to 84) with 1 risk factor; 80% (95% CI 70 to 85) with 2 risk factors; 80% (95% CI 68 to 86) in patient with more than 3 risk factors.

### Sensitivity Analysis

#### COVID 19 Death related VE

In patient aged < 65 years old, the COVID 19 Death related VE estimate was98% (95% CI 93 to 99) in patients without risk factor; 96% (95% CI 90 to 98) in patients with 1 risk factor; 96% (95% CI 89 to 98) in patient with 2 risk factors; and 88% (95% CI 70 to 96) In patient with 3 risk factors and more. In patient aged 1Z65 years old the COVID 19 Death related VE estimate was 93% (95% CI 85 to 97) without risk factor patient; 89% (95% CI 81 to 93) in patients with 1 risk factor;89% (95% CI 79 to 94) in patients with 2 risk factors; and 68% (95% CI 44 to 82) in patients with 3 risk factors and more (Figure 5).

**Figure 5:**
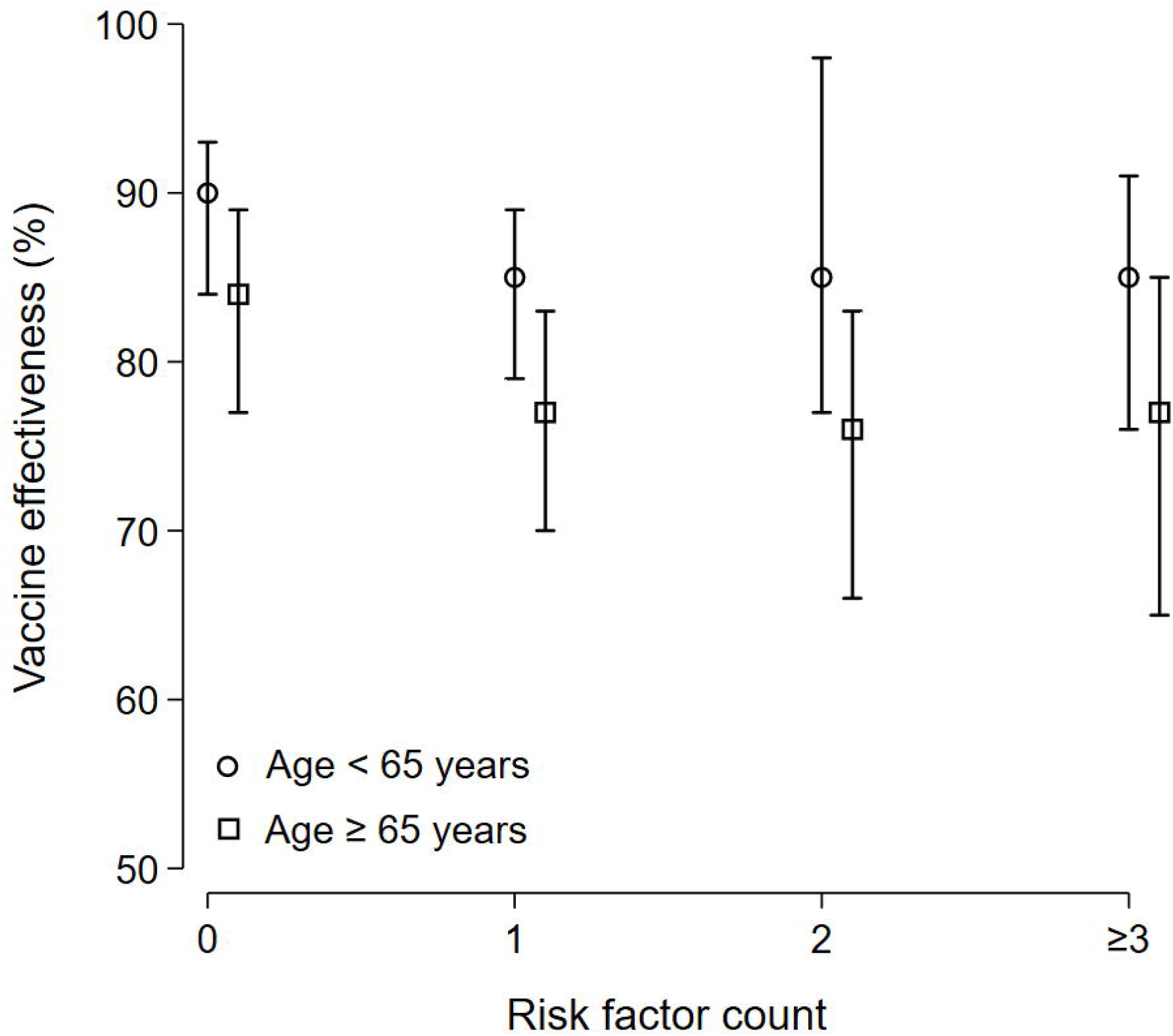
Death related VE : subgroup analysis. In patient aged < 65 years old, the COVID 19 Death related VE estimate was 98% (95% CI 93 to 99) in patients without risk factor; 96% (95% CI 90 to 98) in patients with 1 risk factor; 96% (95% CI 89 to 98) in patient with 2 risk factors; and 88% (95% CI 70 to 96) In patient with 3 risk factors and more. In patient aged lll 65 years old the COVID 19 Death related VE estimate was 93% (95% CI 85 to 97) without risk factor patient; 89% (95% CI 81 to 93) in patients with 1 risk factor; 89% (95% CI 79 to 94) in patients with 2 risk factors; and 68% (95% CI 44 to 82) in patients with 3 risk factors and more.

#### COVID 19 Severe critical hospitalization VE

In patient aged < 65 years old the COVID 19 Severe critical hospitalization related VE estimate was 90% (95% CI 84 to 93) in patients without risk factor; 85% (95% CI 79 to 89) in patients with 1 risk factor; 85% (95% CI 77 to 98) in patients with 2 risk factors; and 85% (95% CI 76 to 91) in patients with 3 risk factors and more. In patient aged 1Z65 years old, the COVID 19 Severe critical hospitalization related VE estimate was 84% (95% CI 77 to 89) In patient without risk factor; 77% (95% CI 70 to 83) in patient with1 risk factor;76% (95% CI 66 to 83) in patient with 2 risk factors; and 77% (95% CI 65 to 85) in patient with 3 risk factors and more (figure 6).

**Figure 6:**
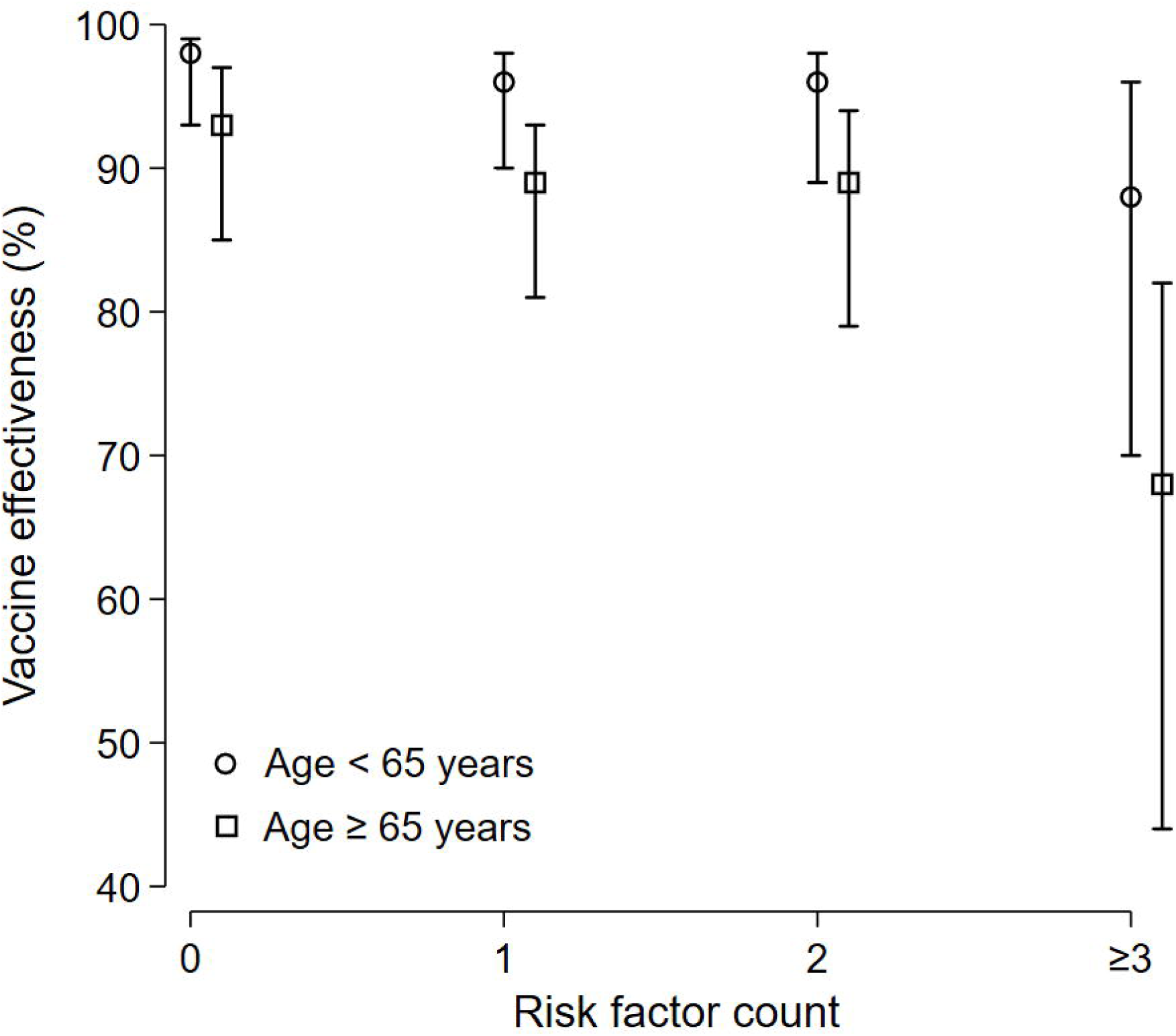
COVID 19 Severe critical hospitalization related VE: subgroup analysis. In patient aged < 65 years old the COVID 19 Severe critical hospitalization related VE estimate was 90% (95% CI 84 to 93) in patients without risk factor; 85% (95% CI 79 to 89) in patients with 1 risk factor; 85% (95% CI 77 to 98) in patients with 2 risk factors; and 85% (95% CI 76 to 91) in patients with 3 risk factors and more. In patient aged lll 65 years old, the COVID 19 Severe critical hospitalization related VE estimate was 84% (95% CI 77 to 89) In patient without risk factor; 77% (95% CI 70 to 83) in patient with1 risk factor; 76% (95% CI 66 to 83) in patient with 2 risk factors; and 77% (95% CI 65 to 85) in patient with 3 risk factors and more.

## Discussion

We present estimates of vaccine effectiveness (VE) for the third booster of the Sinopharm vaccine within the framework of a nationwide mass vaccination campaign aimed at preventing severe and critical cases of COVID-19 hospitalization in the Kingdom of Morocco, with a specific focus on the Omicron VOC variant. Employing a screening methodology, we have ascertained that the administration of the Sinopharm vaccine booster significantly reduces the risk of death (89%) and lowers the risk of severe critical hospitalization (81%) due to COVID-19. This risk reduction is less pronounced in individuals with immune vulnerability associated with advanced age and comorbidities.

In individuals younger than 65 years old without any risk factors, there is a 90% reduction in the risk of severe or critical hospitalization. However, in older patients with one or more risk factors, this reduction decreases to 77%. The risk of COVID-19-related death is reduced by 98% in patients aged under 65 years old without any risk factors. Nevertheless, in older patients with three or more risk factors, this reduction in risk decreases to 68%.

While the effectiveness of COVID-19 vaccines has been extensively evaluated, there is limited data on how clinical risk factors impact vaccine effectiveness, especially in terms of its effectiveness against severe disease. Understanding the variations in vaccine effectiveness among these groups is crucial for guiding decision-makers in formulating strategies for vaccine and antiviral prioritization.

In conclusion, our study demonstrates that individuals who received their booster dose of the inactivated vaccine within 15 to 120 days before hospitalization due to severe or critical manifestations of COVID-19, as well as COVID-19-related deaths, exhibit vaccine effectiveness that aligns with the established guidelines provided by the World Health Organization (WHO). These guidelines specify that vaccine effectiveness against hospitalizations or mortality should meet a minimum threshold of 90%, accompanied by a lower 95% confidence interval of no less than 70%.

It’s important to note that this level of vaccine effectiveness (VE) is not attained in patients aged over 65 who also have comorbidities. This emphasizes the necessity for targeted interventions and ongoing research to tackle the distinctive challenges encountered by this particular population group in achieving optimal vaccine protection. As we further enhance our comprehension of vaccine effictiveness across diverse demographics, these findings offer valuable insights for shaping public health strategies and vaccine distribution initiatives. In conjunction with the vaccination, it is imperative to consider policies related to interventions for vulnerable populations. These policies should encompass strengthening non-pharmaceutical interventions, such as mandatory face mask usage, the use of hand sanitizers and handwashing, as well as social distancing measures. Additionally, optimizing and prioritizing antiviral therapies should be emphasized.

Urquidi et al. demonstrated a positive correlation between full primary vaccination and consistent facemask usage, emphasizing the additional impact of facemasks in augmenting vaccine effectiveness in mitigating severe disease during periods of heightened viral circulation (13,14).

### Future Directions and Conclusions

The timeliness of this research represents a significant strength, as it offers valuable insights into the most recent developments in the battle against COVID-19. This study tackles a highly pertinent and current issue—the efficacy of booster doses of the BBIBP-CorV (Vero Cells) Sinopharm vaccine against the Omicron variant of SARS-CoV-2. By incorporating data from Morocco and focusing on the Sinopharm vaccine’s effectiveness against the Omicron variant of SARS-CoV-2, this research provides a more comprehensive international perspective. The knowledge gleaned from this study may hold implications for other countries contending with similar challenges posed by the Omicron variant.

The effectiveness of Sinopharm boosters in augmenting protection against Omicron variant-related COVID-19 deaths and severe critical hospitalizations is evident. However, this protection diminishes with increasing age and higher risk factors. These findings underscore the significance of tailored vaccination strategies for diverse demographic groups and highlight the protective advantages of administering a third Sinopharm vaccine booster.

It is crucial to note that vaccination remains an effective measure for reducing COVID-19 hospitalizations and fatalities. Nonetheless, other measures, such as face masks and physical distancing, may continue to be necessary for long-term infection control. Our findings offer valuable insights for clinicians, public health policymakers, and researchers concerning the long-term effectiveness of COVID-19 vaccines. These insights can inform clinical and policy recommendations, including considerations regarding the timing of future booster doses.

## Data Availability

The datasets generated for this study are available on request to the corresponding author.

## Author Contributions

JB and RA contributed to the research design, the data collection, the data analysis and data interpretation. JB wrote, and edited the manuscript. RA led the overall study, AM and HM contributed to the data collection. All authors read, and approved the final manuscript.

## Acknowledgments

We thank to the Ministry of Health and Social Protection of the Kingdom of Morocco for providing us with the essential database for our research. We also thank To (“Academie Hassan II des Sciences et Techniques” as part of Morocco consortium for biomedical research on COVID-19) MCBR-Covid619/2021-2023) for their support.

## Conflict of Interest Statement

The authors declare that the research was conducted in the absence of any commercial or financial relationships that could be construed as a potential conflict of interest.

## Notes

### Competing Interest Statement

The authors have declared no competing interest.

### Clinical Protocols

https://osf.io/at3yf

### Author Declarations

Ethics committee/IRB of ethic committee review board for biomedical research at Mohammed V University gave ethical approval for this work

### Summary of Updates

author's names correction

## References

1. Tableau de bord de l’OMS. (2022). https://covid19.who.int/. (Accessed July 30, 2023).

2. Lauring AS, Hodcroft EB. Genetic variants of SARS-CoV-2-what do they mean? JAMA. 2021; 325(6):529–531. PMID: 33404586..

3. Wolter N, Jassat W, Walaza S, Welch R, Moultrie H, Groome M, et al. Early assessment of the clinical severity of the SARS-CoV-2 omicron variant in South Africa: a data linkage study. Lancet. 2022; 399(10323):437–446. PMID: 35065011.

4. Wu N, Joyal-Desmarais K, Ribeiro PAB, Vieira AM, Stojanovic J, Sanuade C, Yip D, Bacon SL. Long-termeffectiveness of COVID-19 vaccines against infections, hospitalisations, and mortality in adults: findings from a rapid living systematic evidence synthesis and meta-analysis up to December, 2022. Lancet Respir Med. 2023 May; 11(5):439-452. doi: 10.1016/S2213-2600(23)00015-2. Epub 2023 Feb 10. PMID: 36780914; PMCID: PMC9917454..

5. Huang, Z., Xu, S., Liu, J. et al. Effectiveness of inactivated COVID-19 vaccines among older adults in Shanghai: retrospective cohort study. Nat Commun 14, 2009 (2023). 10.1038/s41467-023-37673-9

6. Belayachi J, Obtel M, Mhayi A, Razine R, Abouqal R. Long term effectiveness of inactivated vaccine BBIBP-CorV (VeroCells) against COVID-19 associated severe and critical hospitalization in Morocco. PLoS One. 2022 Dec7;17(12):e0278546. doi: 10.1371/journal.pone.0278546. PMID: 36477077; PMCID: PMC9728886.

7. Huang Z, Xu S, Liu J, Wu L, Qiu J, Wang N, Ren J, Li Z, Guo X, Tao F, Chen J, Lu D, Wang Y, Li J, Sun X, Wang W. Effectiveness of inactivated COVID-19 vaccines among older adults in Shanghai: retrospective cohortstudy. Nat Commun. 2023 Apr10;14(1):2009. doi: 10.1038/s41467-023-37673-9. PMID: 37037803; PMCID: PMC10086022.

8. Nittayasoot N, Suphanchaimat R, Thammawijaya P, Jiraphongsa C, Siraprapasiri T, Ploddi K, Pittayawonganon C, Mahasirimongkol S, Tharmaphornpilas P. Real-World Effectiveness of COVID-19 Vaccines againstSevereOutcomesduring the Period of Omicron Predominance in Thailand: A Test-Negative Nationwide Case-Control Study. Vaccines (Basel). 2022 Dec12;10(12):2123. doi: 10.3390/vaccines10122123. PMID: 36560533; PMCID: PMC9785674.

9. Farrington CP. Estimation of vaccine effectiveness using the screening method. Int J Epidemiol. 1993 Aug;22(4):742–6. doi: 10.1093/ije/22.4.742. PMID: 8225751.

10. Guidance on conducting vaccine effectiveness evaluations in the setting of new SARS-CoV-2 variants: Interim guidance, 22 July 2021. Addendum to Evaluation of COVID-19 vaccine effectiveness. https://www.who.int/publications-detail-redirect/WHO-2019-nCoV-vaccine_effectiveness-variants-2021.1. (Accessed July 30, 2023).

11. World Health Organization. COVID-19 clinical management: living guidance https://www.who.int/publications/i/item/WHO-2019-nCoV-clinical-2021-1. (Accessed July 30, 2023).

12. Cohen AL, Taylor T Jr, Farley MM, Schaffner W, Lesher LJ, Gershman KA, Bennett NM, Reingold A, Thomas A, Baumbach J, Harrison LH, Petit S, Beall B, Zell E, Moore M. An assessment of the screening method to evaluate vaccine effectiveness: the case of 7-valent pneumococcalconjugate vaccine in the United States. PLoS One. 2012;7(8):e41785. doi: 10.1371/journal.pone.0041785. Epub 2012 Aug 1. PMID: 22870248; PMCID: PMC3411566.

13. Urquidi C, Santelices E, Lagomarcino AJ, Teresa Valenzuela M, Larrañaga N, Gonzalez E, Pavez A, Wosiack A, Maturana M, Moller P, Pablo Torres J, Muñoz S, O’Ryan G M. The added effect of non-pharmaceutical interventions and lifestyle behaviors on vaccine effectiveness against severe COVID-19 in Chile: A matched case-double control study. Vaccine. 2023 May 2;41(18):2947–2955. doi: 10.1016/j.vaccine.2023.03.060. Epub 2023 Apr 3. PMID: 37024408; PMCID: PMC10067460.

14. Ge Y, Zhang WB, Wu X, Ruktanonchai CW, Liu H, Wang J, Song Y, Liu M, Yan W, Yang J, Cleary E, Qader SH, Atuhaire F, Ruktanonchai NW, Tatem AJ, Lai S. Untangling the changing impact of non-pharmaceutical interventions and vaccination on European COVID-19 trajectories. Nat Commun. 2022 Jun 3;13(1):3106. doi: 10.1038/s41467-022-30897-1. PMID: 35661759; PMCID: PMC9166696..

